# Soluble TREM2 mediates earliest amyloid-associated p-tau increases and cerebral glucose hypermetabolism in Alzheimer’s disease

**DOI:** 10.1101/2022.08.26.22279269

**Authors:** Davina Biel, Marc Suárez-Calvet, Paul Hager, Anna Rubinski, Anna Dewenter, Anna Steward, Sebastian Roemer, Michael Ewers, Christian Haass, Matthias Brendel, Nicolai Franzmeier, for the Alzheimer’s Disease Neuroimaging Initiative (ADNI)

## Abstract

**Background:** Microglial activation occurs early in Alzheimer’s disease (AD) and previous studies reported both detrimental and protective effects of microglia on AD progression. Therefore, it is critical to investigate at which AD stages microglial activation could be protective or detrimental to evaluate microglia as a treatment target. To address this, we used CSF sTREM2 (i.e. Triggering receptor expressed on myeloid cells 2) to investigate disease stage-dependent drivers of microglial activation and to determine downstream consequences on AD biomarker progression.

**Methods:** We included 402 cognitively normal and mild cognitively impaired patients with CSF sTREM2 assessments. To assess AD severity, we included measures of earliest beta-amyloid (i.e. Aβ) in CSF (i.e. Aβ_1-42_) and late-stage fibrillary Aβ pathology (i.e. amyloid-PET centiloid), as well as p-tau_181_ and FDG-PET for assessing downstream changes in tau and cerebral glucose metabolism. To determine disease stage, we stratified participants according to earliest Aβ abnormalities (i.e. Aβ CSF+/PET−; early Aβ-accumulators, n=70) or fully developed fibrillary Aβ pathology (i.e. Aβ CSF+/PET+; late Aβ-accumulators, n=201) plus 131 healthy controls (i.e. Aβ CSF−/PET−).

**Results:** In early Aβ-accumulators, higher centiloid was associated with cross-sectional/longitudinal sTREM2 and p-tau increases, suggesting reactive microglial and p-tau increases in response to earliest Aβ fibrillization. Further, higher sTREM2 mediated the association between centiloid and cross-sectional/longitudinal p-tau increases and higher sTREM2 was associated with FDG-PET hypermetabolism in line with previous findings of increased glucose consumption of activated microglia. In late Aβ-accumulators, we found no association between centiloid and sTREM2 but a cross-sectional association between higher sTREM2, higher p-tau and glucose hypometabolism, suggesting that sTREM2 parallels tau and neurodegeneration rather than Aβ once fully developed Aβ pathology is present.

**Conclusions:** Our findings suggest that sTREM2-related microglial activation occurs in response to earliest Aβ fibrillization, manifests in inflammatory glucose hypermetabolism and may facilitate subsequent p-tau increases in earliest AD, while previous reports of protective sTREM2 effects may occur in later AD stages.

## Background

Alzheimer’s disease (AD) is characterized by the accumulation of beta-amyloid (Aβ), tau, microglial activation, metabolic brain changes, neurodegeneration, and cognitive decline (1). According to the amyloid cascade hypothesis, Aβ accumulation triggers the subsequent development and spreading of hyperphosphorylated tau aggregates which in turn drives neurodegeneration, metabolic decline, and dementia (2–5). Previously it was shown that Aβ-related increases in soluble hyperphosphorylated tau (i.e. p-tau), detectable in plasma (6, 7) and cerebrospinal fluid (CSF) (8), precede tau aggregation. We have shown recently that soluble p-tau increases may in fact drive tau aggregation and spread across interconnected brain regions (9) therefore, soluble p-tau increases may be a key link between Aβ deposition and tau aggregation in AD. However, the underlying mechanisms that link Aβ and subsequent increases in soluble p-tau in CSF or plasma are not well understood.

Here, activation of microglia, the brains innate immune system, may play a key role in modulating these initial events in the amyloid cascade (10). Yet, previous studies have yielded conflicting findings on a detrimental or protective role of microglial activation in AD. For instance, recent in vitro studies reported that activated microglia can induce tau hyperphosphorylation and spread (11) and that activated microglia can release tau seeds which can induce tau aggregation (12). Similarly, studies in sporadic AD patients found that microglial activation is strongly correlated with fluid p-tau but not with Aβ levels (13) and that microglial activation may promote the development of aggregated tau pathology in AD, as measured via tau-PET (10, 14). Indeed, a recent post mortem study investigating the mediating effect of microglial activation on the Aβ to tau association in brain tissue revealed a mediation effect of 33% of microglia for the relationship between Aβ and tau (15). This suggests that microglial activation may be associated with tau hyperphosphorylation and therefore contribute to the development of tau pathology in AD. In addition, activated microglia have been shown to consume high levels of glucose in AD mouse models and AD patients (16), which may manifest in hypermetabolic brain changes that are observed in early-stage AD, when neurodegeneration and ensuing glucose hypometabolism are not yet apparent (17, 18). Thus, glucose hypermetabolism in early AD may not reflect a compensatory mechanism, as suggested previously, but rather reflect activated microglia and neuroinflammation (19, 20). On the contrary, in symptomatic sporadic AD patients and patients with autosomal dominantly inherited AD, higher microglial activation has been associated with attenuated cognitive decline, amyloid accumulation and neurodegeneration (21–23). This suggests a possible protective effect of chronic microglial activation on neuronal integrity and cognition that occurs once pathologic brain changes are severe enough to result in clinically manifested cognitive deficits. Similarly, an animal model has shown that microglial dysfunction is associated with increased Aβ seeding, further suggesting a protective effect of microglia on Aβ pathology development (24). It is therefore of utmost importance for clinical trials trying to target microglial activation as a disease modifying approach to understand i) what drives microglial activation in AD, ii) whether and when microglial activation is beneficial or detrimental and iii) whether the directionality of microglial effects on AD progression depends on disease stage.

The Triggering Receptor Expressed on Myeloid Cell 2 (TREM2) regulates the change of microglia from a homeostatic state to a disease associated state (25, 26), and is a well-established in vivo proxy for microglial activation in AD (21, 27–30). Thus, we used CSF soluble TREM2 (sTREM2) as an in vivo proxy of microglial activation in a well-characterized sample of AD patients and controls to investigate drivers of microglial activation across early vs. late-stage Aβ accumulation in AD and its effects on the development of downstream p-tau and metabolic brain changes. Specifically, we included data of 402 cognitively normal (CN) and mild cognitive impaired (MCI) participants from the ADNI database with available CSF Aβ_1-42_, p-tau_181_, sTREM2, amyloid-PET, and ^18^F-fluorodeoxyglucose PET (FDG-PET). FDG-PET is well established for assessing cerebral glucose uptake and FDG-PET-assessed hypermetabolism has been previously linked to microglial activation (16). To determine disease stage, we classified patients into Aβ CSF+/PET− (early Aβ-accumulators) and Aβ CSF+/PET+ (late Aβ-accumulators) following a previously established approach that allows stratifying individuals by showing earliest signs of Aβ accumulation (i.e. Aβ CSF+/PET−) vs. showing fully developed amyloid pathology (Aβ CSF+/PET+) (31). Additional 131 participants without evidence of Aβ pathology were included as healthy controls (Aβ CSF−/PET−). A subset of participants had available longitudinal p-tau and sTREM2 assessments, based on which we calculated annual sTREM2 and p-tau change rates. Our specific aims were to assess first, whether earliest signs of Aβ accumulation (i.e. in Aβ CSF+/PET−) are associated with sTREM2-related microglial activation and second, whether this initial Aβ-driven microglial activation facilitates subsequent increases in soluble hyperphosphorylated tau (i.e. p-tau). Third, we assessed whether earliest microglial activation is reflected in increased FDG-PET-assessed glucose metabolism, given that activated microglia consume large amounts of glucose (16). Here, we expected higher microglial activation to be associated with glucose hypermetabolism in patients with earliest Aβ accumulation, where neurodegeneration is not yet apparent, vs. hypometabolism in chronic AD phases within late Aβ-accumulators.

## Materials and Methods

### Participants

We included 402 participants from the Alzheimer’s Disease Neuroimaging Initiative (ADNI) with available CSF Aβ_1-42_, p-tau_181_, sTREM2, ^18^F-florbetapir/^18^F-florbetaben amyloid-PET, FDG-PET as well as demographics (sex, age, education) and clinical status. Baseline CSF and PET data had to be obtained within a time window of 6 months. Clinical status was classified by ADNI investigators as cognitively normal (CN; Mini Mental State Examination [MMSE]≥ 24, Clinical Dementia Rating [CDR]=0, non-depressed), mild cognitive impairment (MCI; MMSE≥ 24, CDR=0.5, objective memory-impairment on education-adjusted Wechsler Memory Scale II, preserved activities of daily living) or demented (MMSE=20-26, CDR≥ 0.5, NINCDS/ADRDA criteria for probable AD). ADNI inclusion/exclusion criteria can be found at https://adni.loni.usc.edu/wp-content/uploads/2010/09/ADNI_GeneralProceduresManual.pdf. Demented participants were excluded from this study, since we followed a previous approach of disease stage stratification (31), excluding later stages of AD. To determine disease stage, participants were stratified into early or late Aβ-accumulators based on their Aβ CSF and amyloid-PET status (31). Aβ CSF positivity was determined as CSF Aβ_1−42_<976.6 pg/ml (30) and amyloid-PET positivity was determined as ^8^F-florbetapir>1.11 SUVR (32) or ^18^F-florbetaben PET>1.08 SUVR (33). Participants were then grouped as early Aβ-accumulators (Aβ CSF+/PET−; CN/MCI *n*=30/40) and as late Aβ-accumulators (CSF+/PET+; CN/MCI *n*=41/160). Participants which were classified as Aβ CSF−/PET+ were excluded. In addition, Aβ CSF−/PET− participants were included as a healthy reference group. One participant had abnormal Aβ CSF levels (~4000 pg/ml) and was thus excluded, resulting in 131 controls comprising of CN participants only. A subset of participants had available longitudinal p-tau (early/late/controls *n*=20/75/35) and sTREM2 (early/late/controls *n*=21/75/35) assessments, based on which we calculated annual p-tau and sTREM2 change rates. Ethical approval was obtained by ADNI investigators and all study participants provided written informed consent.

### CSF biomarkers

Aβ_1-42_ and p-tau_181_ levels were assessed by the ADNI biomarker core team at the University of Pennsylvania. An electrochemiluminiscence immunoassays Elecsys on a fully automated Elecsys cobas e 601 instrument and a single lot of reagents for each biomarker were used. For the assessment of sTREM2, a previously described ELISA approach was applied (22, 29, 34). sTREM2 data are provided in the ADNI_HAASS_WASHU_LAB.csv file available in the ADNI database (variable “MSD_STREM2CORRECTED”). A detailed description of the methods can be found online (https://ida.loni.usc.edu).

### MRI and PET acquisition and preprocessing

3T structural MRI was obtained by ADNI employing T1-weighted MPRAGE sequences using unified scanning protocols (http://adni.loni.usc.edu/methods/mri-tool/mri-analysis/). Amyloid-PET was recorded 50-70min after ^18^F-florbetapir injection in 4×5min frames or 90-110min after ^18^F-florbetaben injection in 4×5min frames. FDG-PET was recorded 30-60min after ^18^F-flurodeoxyglucose injection in 6×5min frames. To obtain mean images, recorded time frames were motion corrected and averaged (see also http://adni.loni.usc.edu/methods/pet-analysis-method/pet-analysis/). Using the Advanced Normalization Tools (ANTs; (35)) high-dimensional warping algorithm, nonlinear spatial normalization parameters to Montreal Neurological Institute (MNI) space were estimated based on structural skull-stripped T1-weighted images. Amyloid-PET and FDG-PET images were then co-registered to native-space T1-weighted images and subsequently normalized to MNI space by applying the ANTs-derived normalization parameters. Amyloid-PET SUVRs were intensity normalized to the whole cerebellum and FDG-PET SUVRs to the pons. To harmonize between both amyloid-PET tracers, global amyloid-PET SUVRs across ^18^F-florbetapir and ^18^F-florbetaben were transformed to centiloid using equations provided by ADNI (36).

### Statistical analyses

All statistical analyses were computed using R statistical software version 4.0.2 (http://www.R-project.org; 37).

Baseline characteristics between groups were compared using ANOVAs for continuous and chi-squared tests for categorial data.

To test whether microglial activation drives soluble p-tau increases in early Aβ-accumulators, we first assessed cross-sectional disease stage-dependent (i.e. early vs. late Aβ-accumulators) associations between centiloid and sTREM2, and between centiloid and p-tau, using linear regressions. Here, we used sTREM2 or p-tau as the dependent variable, and centiloid as the independent variable. To assess longitudinal disease stage-dependent associations, we first calculated slope estimates for annual change rates of sTREM2 and p-tau for those participants with available longitudinal sTREM2 and p-tau data. Here, linear mixed models were fitted using sTREM2 or p-tau as dependent variable and time (i.e. years from baseline) as independent variable, adjusting for subject-specific random slope and intercept. Subsequently, we performed linear regressions using annual change rates of sTREM2 or p-tau as the dependent variable and baseline centiloid as the independent variable. In addition, analyses were repeated using CSF Aβ instead of centiloid for testing associations between soluble Aβ and sTREM2 or p-tau. The regression models were controlled for age, sex, education, and clinical status (i.e. CN or MCI). The models testing the control group were not controlled for clinical status since the group only comprised of CN participants. Finally, bootstrapped mediation analyses using 1000 iterations were applied using the mediation package in R (https://cran.r-project.org/web/packages/mediation/mediation.pdf) for assessing cross-sectional and longitudinal associations, using centiloid as the predictor variable, p-tau (i.e. cross-sectional) or p-tau change rate (i.e. longitudinal) as the dependent variable, and sTREM2 as the mediator. Subsequently, we tested the reverse associations (i.e. p-tau as the mediator of centiloid’s effect on sTREM2). All mediation analyses were controlled for age, sex, education, and clinical status.

In the next step, we assessed whether sTREM2-related microglial activation manifests in glucose hypermetabolism in early AD stages when neurodegeneration is typically not yet apparent (i.e. Aβ CSF+/PET−) vs. glucose hypometabolism in later disease stage (i.e. Aβ CSF+/PET+). To this end, we first calculated FDG-PET z-scores by referencing FDG-PET signals to 131 CN Aβ− controls. We then tested for an sTREM2 by group (i.e. Aβ CSF+/PET− vs. Aβ CSF+/PET+) interaction on FDG-PET signal in a meta-ROI that typically captures AD-related glucose metabolism changes (38). Next, we repeated the analysis in a brain wide manner across 200 ROIs that cover the entire neocortex (39), in order to map the pattern of microglial activation effects on glucose hyper-vs. hypometabolism depending on disease stage. To that end, we computed linear regressions for the associations between sTREM2 and each of the 200 FDG-PET ROIs. To test whether early/late Aβ-accumulators differ regarding sTREM2-related glucose metabolism, T-values of the association between sTREM2 and FDG-PET were compared between groups using t-tests. The models were controlled for age, sex, education, and clinical status.

## Results

We stratified participants by evidence for early-vs. late-stage Aβ pathology, using a previously established approach that combines CSF assessments of soluble Aβ_1-42_ and amyloid-PET assessments of fibrillar Aβ (31). CSF Aβ abnormality is assumed to reflect early Aβ oligomerization and precedes amyloid-PET positivity reflecting mostly fibrillary forms of Aβ (40). Therefore, we grouped participants into i) early Aβ-accumulators (i.e. Aβ CSF+/PET−; *n*=70 participants, CN=30; MCI=40) with evidence for reduced Aβ_1-42_ in CSF but no suprathreshold fibrillar Aβ pathology on PET vs. ii) late Aβ-accumulators (i.e. Aβ CSF+/PET+; *n*=201, CN=41; MCI=160), with evidence for abnormal Aβ in both CSF and PET. An additional pool of 131 cognitively normal subjects without evidence of abnormal Aβ on either CSF or PET was included as a control group. Longitudinal CSF data were available for a subset of participants for sTREM2 (early Aβ/late Aβ/controls *n*=21/75/35) and p-tau (early Aβ/late Aβ/controls *n*=20/75/35) with an average follow-up time from baseline CSF assessment of 1.99±0.09 years. Descriptive baseline statistics stratified by groups are shown in **Table 1**.

**Table 1.** Demographic and clinical data stratified by group.

| | Controls<br>(Aβ CSF-/PET-) | Early Aβ-accumulators<br>(Aβ CSF+/PET-) | Late Aβ-accumulators<br>(Aβ CSF+/PET+) | p-value |
| --- | --- | --- | --- | --- |
| Cross-sectional |  |  |  |  |
| *N* | 131 | 70 | 201 |  |
| Diagnostic (CN/MCI) | 131/0 | 30/40 | 41/160 | <0.001 |
| Sex (male/female) | 64/67 | 47/23 | 112/89 | 0.045 |
| Age in years | 72.67 (6.39) | 71.65 (7.68) | 73.60 (6.53) | 0.093 |
| Years of education | 16.85 (2.43)<sup>c</sup> | 17.13 (2.24)<sup>c</sup> | 16.00 (2.74)<sup>a,b</sup> | <0.001 |
| CSF Aβ (pg/ml) | 1553.122 (281.109)<sup>b,c</sup> | 777.819 (147.169)<sup>a,c</sup> | 656.42 (174.79)<sup>a,b</sup> | <0.001 |
| CSF p-tau (pg/ml) | 20.241 (7.059)<sup>b,c</sup> | 15.825 (7.493)<sup>a,c</sup> | 32.807 (14.780)<sup>a,b</sup> | <0.001 |
| CSF sTREM2 (pg/ml) | 4099.933 (1980.197)<sup>b</sup> | 2955.118 (1751.746)<sup>a,c</sup> | 3984.107 (2194.033)<sup>b</sup> | <0.001 |
| Amyloid-PET (centiloid) | -9.0315 (12.656)<sup>c</sup> | -2.901 (13.530)<sup>c</sup> | 78.228 (33.871)<sup>a,b</sup> | <0.001 |
| FDG-PET global z-score | - | -0.132 (0.649) | -0.251 (0.532) | 0.13 |
| FDG-PET meta-ROI z-score | - | -0.214 (0.826)<sup>c</sup> | -0.546 (0.723)<sup>b</sup> | 0.002 |
| Longitudinal |  |  |  |  |
| *N* | 35 | 20 | 75 |  |
| Follow-up CSF p-tau (mean years) | 1.96 (0.13)<sup>b</sup> | 2.05 (0.08)<sup>a,c</sup> | 1.99 (0.07)<sup>b</sup> | 0.002 |
| *N* | 35 | 21 | 75 |  |
| Follow-up CSF sTREM2 (mean years) | 1.96 (0.13)<sup>b</sup> | 2.05 (0.08)<sup>a</sup> | 1.99 (0.07) | 0.004 |

Values are presented as mean (SD); p-values were derived from ANOVAs for continuous measures and from Chi-squared tests for categorical measures.
Mean values significantly ( $p < 0.05$ , Post-hoc tests) different from—
a Controls
b Early Aβ-accumulators
c Late Aβ-accumulators

### Early but not late-stage Aβ accumulation is associated with higher sTREM2

We tested first whether evidence for earliest Aβ abnormality in CSF but not yet in PET (i.e. early Aβ accumulators) is associated with an sTREM2-related microglial response and p-tau increases. To this end, we used linear regression to determine the association between amyloid-PET as a marker of fibrillary Aβ pathology (i.e. centiloid) and sTREM2 in early Aβ-accumulators. Here, higher centiloid at baseline was associated with higher cross-sectional p-tau (β=0.259, T=2.200, *p*=0.032; **Fig.1A**, top panel) and higher sTREM2 (β=0.254, T=2.268, *p*=0.027; **Fig.1B**, top panel). Further, higher sTREM2 levels were associated with higher p-tau levels (β=0.587, T=5.425, *p*<0.001). We obtained congruent results using longitudinal CSF data, showing that higher centiloid at baseline was associated with faster subsequent change rates in in p-tau (β=0.550, T=2.975, *p*=0.010; **Fig.1C**, top panel) and faster changes in sTREM2 (β=0.535, T=3.725, *p*=0.002; **Fig.1D**, top panel). Using bootstrapped mediation analyses with 1000 iterations, we additionally found that the association between higher centiloid and higher p-tau was fully mediated by sTREM2 in early Aβ accumulators, both for cross-sectional p-tau (average causal mediation effect [ACME]: B=0.133, 95%CI=[0.0039-0.27], *p*=0.038; **Fig.1E**, top panel) as well subsequent p-tau change rates (ACME: B=0.450, 95%CI=0.1352-0.82, *p*=0.004; **Fig.1F**, top panel). Testing the reverse mediation models, i.e. whether p-tau mediates the effect of centiloid on sTREM2 yielded a similar mediation effect for cross-sectional sTREM2 levels (ACME: B=0.132, CI=0.0084-0.26, *p*=0.030) but a much lower mediation effect for subsequent sTREM2 change rates in longitudinal analyses (ACME: B=0.273, CI=0.0437-0.55, *p*=0.014). These results suggest that earliest Aβ accumulation may induce reactive sTREM2-related microglial activation which may in turn facilitate p-tau increases.

**Figure 1.**
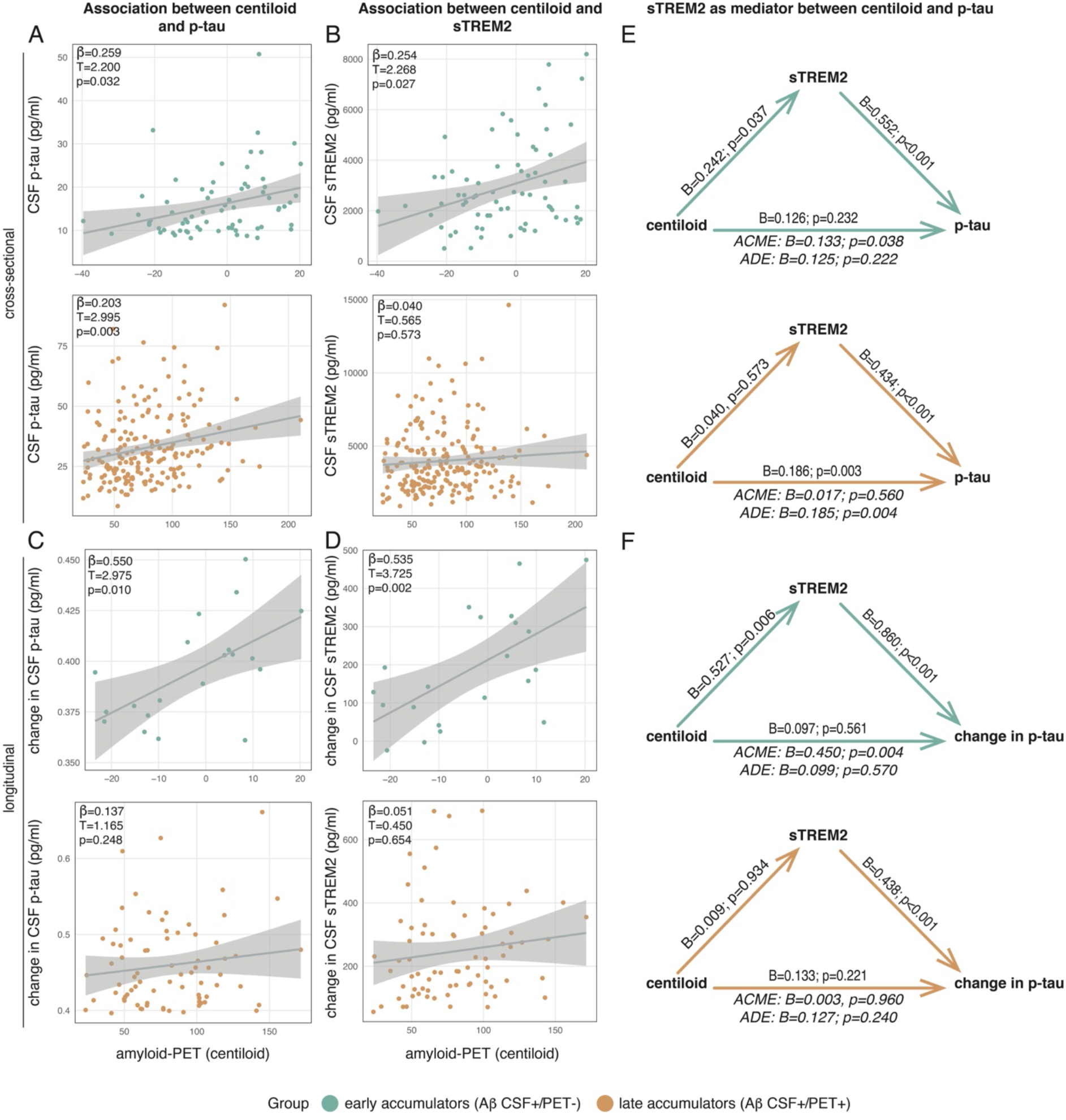
Cross-sectional and longitudinal analysis of the association between amyloid-PET (in centiloid), CSF p-tau, and CSF sTREM2 in early Aβ-accumulators (i.e. Aβ CSF+/PET−) and late Aβ-accumulators (i.e. Aβ CSF+/PET+). Cross-sectional associations between **A)** centiloid and p-tau and **B)** centiloid and sTREM2. Longitudinal associations between **C)** centiloid and change in p-tau and **D)** centiloid and change in sTREM2. Standardized beta-estimates (β), T-values, and p-values were derived from linear regressions controlling for age, sex, education, and clinical status. **E)** Cross-sectional mediation analyses with centiloid as predictor, sTREM2 as mediator, and p-tau as dependent variable. **F)** Longitudinal mediation analyses with centiloid as predictor, sTREM2 as mediator, and change in p-tau as dependent variable. Early Aβ-accumulators are displayed in green, and late Aβ-accumulators in orange. Beta-estimates (B) and p-values for each path are displayed on the respective arrow. The average causal mediation effect [ACME] and the average direct effect [ADE] are displayed under each mediation triangle. All models are controlled for age, sex, education, and clinical status.

When assessing the above-described analyses in late Aβ-accumulators, no association was found between centiloid and sTREM2, neither for cross-sectional sTREM2 (β=0.040, T=0.565, *p*=0.573; **Fig.1B**, bottom panel) nor for longitudinal sTREM2 change rates (β=0.051, T=0.450, *p*=0.654; **Fig.1D**, bottom panel). This suggests that sTREM2 increases are no longer driven by Aβ once fully developed fibrillar Aβ pathology is present. Yet, there was an association between higher baseline sTREM2, higher baseline p-tau (β=0.441, T=7.004, *p*<0.001) and longitudinal changes in p-tau (β=0.439, T=3.777, *p*<0.001), suggesting that sTREM2 is more strongly coupled to p-tau in late Aβ-accumulators. Higher centiloid was associated with higher cross-sectional p-tau (β=0.203, T=2.995, *p*=0.003; **Fig.1A**, bottom panel), but no association was found between baseline centiloid and subsequent p-tau change rates (*n*=75, β=0.137, T=1.165, *p*=0.248; **Fig.1C**, bottom panel). Given that there was no association between centiloid and sTREM2 in late Aβ-accumulators, we did not detect a mediation effect of sTREM2 for the association between centiloid and p-tau, neither cross-sectionally (ACME: B=0.017, CI=−0.0444-0.08, *p*=0.560; **Fig.1E**, bottom panel), nor longitudinally (ACME: B=0.003, CI=−0.108-0.10, *p*=0.960; **Fig.1F**, bottom panel). Together, these results suggest that sTREM2 dynamics are more closely associated with p-tau dynamics rather than fibrillar Aβ in patients with fully developed Aβ pathology as indicated by reduced CSF Aβ as well as a positive amyloid-PET scan.

When performing the same analyses in Aβ CSF−/PET− healthy controls, no significant cross-sectional or longitudinal associations between centiloid and sTREM2 (cross-sectional: β=0.118, T=1.273, *p*=0.205; longitudinal: β=−0.049, T=−0.240, *p*=0.812) or p-tau (cross-sectional: β=0.091, T=0.967, *p*=0.335; longitudinal: β=−0.119, T=−0.582, *p*=0.565) were observed, suggesting that the association between Aβ fibrillization and sTREM2 increases are specific for patients who show earliest signs of AD pathophysiology as indicated by reduced CSF Aβ_1-42_ levels. Together, these findings suggest that earliest fibrillization of Aβ is associated with reactive sTREM2 increases, which may in turn facilitate increases in p-tau in early Aβ−accumulators, while sTREM2 dynamics may uncouple from the extent of fibrillar Aβ pathology at later stages but rather parallel p-tau increases.

Lastly, the above-described analyses were repeated using CSF Aβ_1−42_ instead of centiloid for testing associations between soluble Aβ and p-tau or sTREM2. Except for an association between CSF Aβ_1−42_ and p-tau in late accumulators (*p*=0.004), no associations were observed (**Table 2**), which is in accordance with previous work (30). This suggests that sTREM2 increases are specifically associated with fibrillization of Aβ as assessed via amyloid-PET.

**Table 2.** Associations between CSF Aβ_1-42_ and p-tau or sTREM2. The table displays standardized beta-estimates (β), T-values, and p-values. The regression models are controlled for age, sex, education, and clinical status.

|  | Cross-sectional |  |  | Longitudinal |  |  |
| --- | --- | --- | --- | --- | --- | --- |
| | $\beta$ | T | p | $\beta$ | T | p |
| *Early Aβ-accumulators (Aβ CSF+/PET-)* |  |  |  |  |  |  |
| p-tau ~ A$\beta_{1-42}$ | -0.225 | -1.911 | 0.061 | -0.215 | -0.863 | 0.403 |
| sTREM2 ~ A$\beta_{1-42}$ | 0.128 | 1.124 | 0.265 | -0.006 | -0.029 | 0.978 |
| *Late Aβ-accumulators (Aβ CSF+/PET+)* |  |  |  |  |  |  |
| p-tau ~ A$\beta_{1-42}$ | 0.196 | 2.894 | 0.004 | 0.094 | 0.791 | 0.432 |
| sTREM2 ~ A$\beta_{1-42}$ | 0.069 | 0.979 | 0.329 | 0.134 | 1.197 | 0.236 |

### Microglial activation is reflected in cerebral glucose metabolism across AD stages

Next, we tested whether there is a non-linear relationship between sTREM2 levels and FDG-PET-assessed glucose metabolism across the spectrum of Aβ accumulation compared to controls. Microglia consume large amounts of glucose (16), hence sTREM2-related microglial activation may result in FDG-PET hypermetabolism in the earliest stages of Aβ accumulation, where neurodegeneration is typically not yet apparent. In late Aβ accumulators, sTREM2-related microglial activation parallels p-tau which has been associated with subsequent neurodegeneration (9, 41). Hence higher sTREM2 may be associated with FDG-PET hypometabolism in subjects with late-stage Aβ accumulation. To test this, we summarized FDG-PET of early and late Aβ-accumulators across AD vulnerable brain regions (38) and referenced the mean FDG-PET signal to 131 controls (i.e. Aβ CSF−/PET−) to derive FDG-PET z-scores that allow to determine whether FDG-PET metabolism is higher or lower than in a reference group of healthy controls. Using linear regression, we observed a significant sTREM2 by group interaction on FDG-PET meta-ROI z-scores (β=-0.378, T=−1.980, *p*=0.049; **Fig.2A**, controlled for age, sex, education, and clinical status), showing that sTREM2 was associated with higher FDG-PET in early Aβ-accumulators vs. lower FDG-PET in late Aβ-accumulators. To map the spatial pattern of brain regions in which higher sTREM2 is associated with higher FDG-PET in early Aβ-accumulators, vs. lower FDG-PET uptake in late Aβ-accumulators, we assessed the association between sTREM2 and FDG-PET z-scores for early and late Aβ-accumulators separately across 200 cortical brain regions included in the Schaefer brain atlas (39) using linear regression models controlling for age, sex, education, and clinical status. When projecting the T-values of the association between sTREM2 levels and FDG-PET to the brain surface (**Fig.2B**, left panel), we found consistent and brain-wide positive associations between sTREM2 and FDG-PET for early Aβ-accumulators, vs. consistent negative associations between sTREM2 and FDG-PET for late Aβ-accumulators. Comparing the T-value distributions of the association between sTREM2 levels and FDG-PET between early and late Aβ-accumulators showed a significant group difference (T=35.78, *p*<0.001, **Fig.2B**, right panel), supporting the view that there is a non-linear relationship between sTREM2 levels and FDG-PET across the spectrum of Aβ deposition, where sTREM2 is associated with relative FDG-PET hypermetabolism in early Aβ-accumulators vs. relative hypometabolism in late Aβ-accumulators. These findings suggest that elevated glucose metabolism in the early disease stage, before neurodegeneration is present, reflects neuroinflammation while in the later disease stage, higher sTREM2 parallels neuronal loss and tau aggregation, and therefore may manifest in metabolic decline.

**Figure 2.**
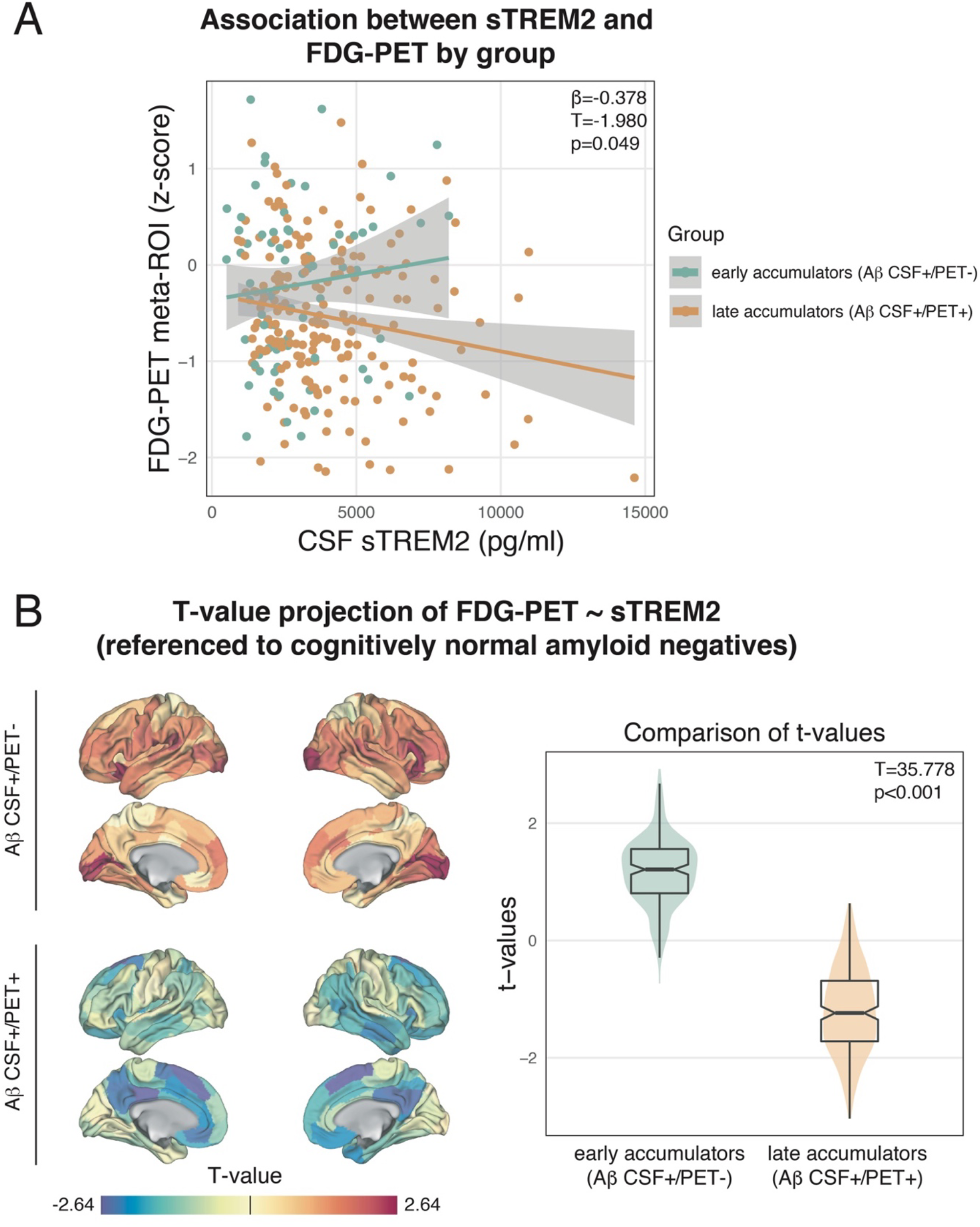
Association between CSF sTREM2 and FDG-PET in early Aβ-accumulators (i.e. Aβ CSF+/PET−) and late Aβ-accumulators (i.e. Aβ CSF+/PET+). **A)** Plot shows significant (p<0.05) sTREM2 by group interaction on FDG-PET z-scores within an FDG-PET meta-ROI (Landau & Jagust, 2011). **B)** T-value projection of the association between sTREM2 on FDG-PET, stratified by group. FDG-PET z-scores were derived by referencing FDG-PET SUVRs to cognitively normal controls (i.e. *n*=131; Aβ CSF−/PET−). The models are controlled for age, sex, education, and clinical status.

## Discussion

In this combined CSF biomarker and neuroimaging study, we systematically assessed the drivers of earliest sTREM2-related microglial activation in AD and its consequences on downstream changes in the amyloid cascade, including soluble p-tau increases and changes in cerebral glucose metabolism. In summary we found that in early Aβ-accumulators, higher Aβ was associated with higher sTREM2 and that sTREM2 mediated the association between earliest PET-assessed fibrillary Aβ deposition and soluble p-tau increases. In contrast, higher sTREM2 was no longer associated with Aβ but paralleled p-tau increases in late Aβ-accumulators, suggesting that sTREM2 is more strongly coupled to the increase in soluble p-tau levels once fully developed fibrillary Aβ pathology is present. Higher sTREM2 levels were further associated with FDG-PET-assessed glucose hypermetabolism in early Aβ-accumulators but with glucose hypometabolism in late Aβ-accumulators. This suggests that increases in glucose metabolism observed in early-stage AD may reflect Aβ-related neuroinflammation rather than a compensatory effect (17, 19). Together, our results suggest that sTREM2-related microglial activation is a key element of the amyloid cascade that may accelerate earliest p-tau increases and metabolic brain changes, while previously reported protective effects of sTREM2-related microglial activation on attenuated neurodegeneration and symptom progression may be observed in later disease stages (21, 22).

Our first finding showed that the earliest sTREM2-related microglial response in AD is associated with fibrillar yet subthreshold Aβ increases (i.e. centiloids below 20), and that higher sTREM2 mediated the earliest Aβ-related increases in soluble p-tau levels. Therefore, sTREM2-related microglial activation may play a key role in the earliest events of the amyloid cascade. In contrast, in participants showing fully developed Aβ pathology as indicated by combined positivity on both CSF and PET (i.e. late Aβ-accumulators), only associations between fibrillar Aβ (i.e. centiloid) and p-tau or between p-tau and sTREM2 were found, suggesting that in late Aβ-accumulators, increases in sTREM2-related microglial activation are no longer driven by the severity of fibrillar Aβ pathology but rather parallel the increases of soluble p-tau. Notably, we and others (30) could not detect any associations between CSF Aβ levels and sTREM2, hence early insoluble but not soluble forms of Aβ might be associated with sTREM2-related microglial activation and subsequent p-tau increases. Importantly, our results remained consistent when repeating the analysis in a subset of participants with available longitudinal CSF data, showing that sTREM2 increases at baseline predict and mediate subsequent Aβ-related increases in p-tau in early but not in late Aβ-accumulators. These findings support the view that earliest Aβ fibrillization induces a sTREM2-related activation of microglia, which is in turn associated with p-tau increases, while sTREM2 increases may uncouple from Aβ severity at later stages. Our findings of microglial activation as a mediator of soluble p-tau increases is in line with a recent post mortem study in older adults showing that activated microglia partly mediated the relationship between Aβ and tau (15). In addition, several preclinical studies showed that activated microglia can enhance tau phosphorylation in animal models of AD and other tauopathies (11, 42–44). Similarly, another study in a rat model of tauopathy could show that animals that are genetically prone to neuroinflammation show stronger neurofibrillary tau pathology compared to animals with less neuroinflammation (45). A further study using brain tissue of AD patients and AD mice could show that microglia phagocytose hyperphosphorylated tau seeds but are incapable of fully neutralizing tau seeding activity and instead release pathological tau seeds into the extracellular space, which can induce a cascade of subsequent tau hyperphosphorylation, misfolding and spread (46). Moreover, a large-scale biomarker study in AD patients with combined amyloid-PET, tau-PET, and TSPO-PET for assessing microglial activation levels could show that elevated microglial activation may promote the aggregation and spreading of fibrillary tau deposits (10). However, there are also contrasting reports, showing that TREM2 loss of function is associated with a drastically elevated AD risk (47, 48) and with facilitated Aβ-associated tau seeding in AD mice expressing both Aβ and tau pathology (49, 50). Further, we reported previously that higher baseline sTREM2 levels in symptomatic AD patients are associated with lower tau-PET levels several years later (21). Conflicting findings of microglial activation on the development of tau pathology may be explained by several factors, including the use of different animal models of AD, which recapitulate different aspects of AD pathophysiology, or by the use of different markers of tau pathology including soluble and fibrillar forms of tau. CSF p-tau reflects hyperphosphorylated tau in its soluble form, one of the earliest tau-related changes in AD that is closely associated with Aβ, preceding the formation of intracellular neurofibrillary tau aggregates (9, 51). Therefore, microglia may have different effects on p-tau hyperphosphorylation and aggregation, which will be important to study in greater detail in future studies by combining fluid and PET biomarkers of microglial activation and tau pathology. Further, our study showed that the inclusion of different AD stages can have a drastic impact on the association between sTREM2 levels and downstream AD biomarkers, which may be a key confounder in previous studies. This is also supported by a previous study using an APPPS1-21 mouse model, showing that TREM2-related microglial activation can have opposing effects on Aβ pathology, depending on disease stage (52). Nevertheless, our findings highlight the view that microglia are crucially involved in the amyloid cascade, yet the directionality of effects may be modulated by disease stage and other pathophysiological events. Therefore, we believe that it is crucial to better understand how microglia and sTREM2 are involved in the molecular progression of tau pathology, from tau hyperphosphorylation and increases in soluble p-tau to the development and spread of fibrillary tau pathology, in order to evaluate microglia as a treatment target.

As a second finding, we report that microglial activation was associated with increased FDG-PET-assessed glucose metabolism in early Aβ-accumulators, and with decreased glucose metabolism in late Aβ-accumulators compared to healthy controls. This observation aligns well with previous work, showing that activated microglia consume high levels of glucose (16) and should thus be reflected in increased FDG-PET. The metabolic drop that we observed in later disease might in contrast reflect neurodegenerative processes and neuronal death (4). Increased glucose metabolism has been previously reported in early AD stages (17, 19) and interpreted as possible compensatory neuronal activity (19), however, proof of an actual advantage of these compensatory mechanisms has been lacking. A match between regional hypermetabolism in early disease stages followed by hypometabolism in later stages has been observed in a previous cross-sectional study (17), where cognitively normal older adults with higher levels of Aβ showed glucose hypermetabolism in those regions, which were most susceptible to AD-related hypometabolism in advanced disease. In addition, in a study including the DIAN cohort, the biggest database of autosomal dominant AD patients, increased FDG-PET signal was detected ~25 years before estimated symptom onset in mutation carriers compared to non-carriers. With advanced disease, hypometabolism was mostly observed in regions, that were previously hypermetabolic (53). However, the authors caution the low number of participants (*n*=11) which were included in the analysis. Importantly, we detected statistically significant associations between sTREM2-related microglial activation and glucose metabolism depending on disease stage. Considering previous work, our findings suggest that different stages of neuroinflammation and neurodegeneration may induce a non-linear trajectory of metabolic brain changes in AD. This is also supported by a recent study showing associations between astrogliosis, i.e. another soluble marker of neuroinflammation, and higher FDG-PET metabolism in earliest AD (54). Future studies using more advanced TSPO-PET for the regional assessment of neuroinflammation should further investigate whether regions affected by high Aβ load show increased signs of neuroinflammation and whether this is coupled by glucose hypermetabolism in earliest AD.

Our findings have important implications for microglial-related treatment strategies. Previous trials focusing on the reduction of Aβ plaques have failed to significantly reduce cognitive dysfunction (55). Thus, treatments that are directly targeting the pathogenesis of tau, which is much closer linked to neurodegeneration and cognitive decline than Aβ (56–61), might obtain better results. In the present study, we showed that in earliest Aβ accumulation microglial activation moderates the association between Aβ and tau, thus, a treatment that targets microglial activation might reduce increases in p-tau and therefore, prevent tau aggregates and following neurodegeneration. Here, the time window for an optimal treatment effect will be critical, since our models showed that microglial activation only mediates p-tau increases in early Aβ-accumulators, while in later disease stages, beneficial effects of activated microglia on cognition could be observed (21, 28). Therefore, future studies are highly needed to reveal the ideal time windows for microglial-related treatment that might target reduction or enhancement of microglia, depending on disease stage.

A clear strength of the present study is its multi-modal design including CSF biomarkers and PET imaging. When interpreting the data, however, several caveats should be addressed. First, we cannot ensure that all participants of the early Aβ-accumulators group will progress to fully developed fibrillar Aβ pathology due to a lack of longitudinal amyloid-PET data. To minimize the risk of wrong allocation, participants showing an unexpected Aβ biomarker pattern (i.e. CSF−/PET+) or those which were already demented were excluded from this study (31). Second, we could not assess p-tau by sTREM2 interactions on tau aggregates since tau-PET data were limited and thus not sufficient to reliably assess relationships in the current dataset. Nevertheless, we encourage future studies to assess, whether sTREM2 in later disease stages shows protective effects on tau aggregates, thereby preventing neuronal death and atrophy which might explain previous findings on protective effects of sTREM2 on neurodegeneration and cognitive decline (22). Finally, for generalization of the results, the analysis should be replicated in diverse cohorts of ethnical variety, since race has been shown to have an effect on markers of microglial activation (62).

## Conclusions

In summary, our findings support disease stage-dependent effects of microglial activation on AD disease progression. Besides previously reported beneficial effects of sTREM2-related microglial activation in advanced AD, we show that in patients with earliest Aβ abnormalities, microglial activation is associated with increases in p-tau and neuroinflammation which is reflected in glucose hypermetabolism. Our findings have important clinical implications as they suggest that targeting enhancement of sTREM2-related microglial activation may have opposing effects on the progression of AD pathophysiology in early vs. later stages of AD.

## Data Availability

All data used in this manuscript are publicly available from the ADNI database (adni.loni.usc.edu) upon registration and compliance with the data use agreement. The data that support the findings of this study are available on reasonable request from the corresponding author.

## Abbreviations

Aβ: amyloid-beta
AD: Alzheimer’s disease
ADNI: Alzheimer’s Disease Neuroimaging Initiative
CN: cognitively normal
CSF: cerebrospinal fluid
FDG: F-flurodeoxyglucose
MCI: mild cognitively impaired
MMSE: Mini Mental State Examination
ROI: region of interest
sTREM2: soluble Triggering Receptor Expressed on Myeloid Cell 2
SUVR: standardized uptake value ratio

## Declarations

### Ethics approval and consent to participate

Ethics approval was obtained by the ADNI investigators from the local ethical committees of all involved sites. The study was conducted in accordance with the Declaration of Helsinki and all study participants provided written informed consent. All work complied with ethical regulations for work with human participants.

### Consent for publication

Not applicable.

### Authors’ contributions

D.B.: study concept and design, data processing, statistical analysis, interpretation of the results, and writing the manuscript. M.S.: critical revision of the manuscript. P.H.: data processing, critical revision of the manuscript. A.R.: critical revision of the manuscript. A.D.: critical revision of the manuscript. A.S.: critical revision of the manuscript. S.R.: critical revision of the manuscript. M.E.: critical revision of the manuscript. C.H.: data acquisition, critical revision of the manuscript. M.B.: study concept and design, interpretation of the results, critical revision of the manuscript. N.F.: study concept and design, data processing, statistical analysis, interpretation of the results, and writing the manuscript. ADNI provided all data used for this study.

## Acknowledgements

We thank the ADNI participants and their families who made this study possible.

## References

1. Jack CR, Bennett DA, Blennow K, Carrillo MC, Dunn B, Haeberlein SB, et al. NIA-AA Research Framework: Toward a biological definition of Alzheimer,Äôs disease. Alzheimer’s & dementia : the journal of the Alzheimer’s Association. 2018;14(4):535–62.

2. Karran E, Mercken M, De Strooper B. The amyloid cascade hypothesis for Alzheimer’s disease: an appraisal for the development of therapeutics. Nat Rev Drug Discov. 2011;10(9):698–712.

3. La Joie R, Visani AV, Baker SL, Brown JA, Bourakova V, Cha J, et al. Prospective longitudinal atrophy in Alzheimer’s disease correlates with the intensity and topography of baseline tau-PET. Sci Transl Med. 2020;12(524).

4. Strom A, Iaccarino L, Edwards L, Lesman-Segev OH, Soleimani-Meigooni DN, Pham J, et al. Cortical hypometabolism reflects local atrophy and tau pathology in symptomatic Alzheimer’s disease. Brain. 2022;145(2):713–28.

5. Haass C, Selkoe D. If amyloid drives Alzheimer disease, why have anti-amyloid therapies not yet slowed cognitive decline? PLoS Biol. 2022;20(7):e3001694.

6. Janelidze S, Berron D, Smith R, Strandberg O, Proctor NK, Dage JL, et al. Associations of Plasma Phospho-Tau217 Levels With Tau Positron Emission Tomography in Early Alzheimer Disease. JAMA Neurol. 2021;78(2):149–56.

7. Moscoso A, Grothe MJ, Ashton NJ, Karikari TK, Rodriguez JL, Snellman A, et al. Time course of phosphorylated-tau181 in blood across the Alzheimer’s disease spectrum. Brain. 2021;144(1):325–39.

8. Mattsson-Carlgren N, Andersson E, Janelidze S, Ossenkoppele R, Insel P, Strandberg O, et al. Abeta deposition is associated with increases in soluble and phosphorylated tau that precede a positive Tau PET in Alzheimer’s disease. Sci Adv. 2020;6(16):eaaz2387.

9. Binette AP, Franzmeier N, Spotorno N, Ewers M, Brendel M, Biel D, et al. Amyloid-associated increases in soluble tau is a key driver in accumulation of tau aggregates and cognitive decline in early Alzheimer. medRxiv. 2022:2022.01.07.22268767.

10. Pascoal TA, Benedet AL, Ashton NJ, Kang MS, Therriault J, Chamoun M, et al. Microglial activation and tau propagate jointly across Braak stages. Nat Med. 2021;27(9):1592–9.

11. Maphis N, Xu G, Kokiko-Cochran ON, Jiang S, Cardona A, Ransohoff RM, et al. Reactive microglia drive tau pathology and contribute to the spreading of pathological tau in the brain. Brain : a journal of neurology. 2015;138(Pt 6):1738–55.

12. Brelstaff JH, Mason M, Katsinelos T, McEwan WA, Ghetti B, Tolkovsky AM, et al. Microglia become hypofunctional and release metalloproteases and tau seeds when phagocytosing live neurons with P301S tau aggregates. Sci Adv. 2021;7(43):eabg4980.

13. Suárez-Calvet M, Morenas-Rodríguez E, Kleinberger G, Schlepckow K, Caballero MÁA, Franzmeier N, et al. Early increase of CSF sTREM2 in Alzheimer’s disease is associated with tau related-neurodegeneration but not with amyloid-β pathology. Molecular Neurodegeneration. 2019;14(1):1.

14. Vogels T, Murgoci AN, Hromadka T. Intersection of pathological tau and microglia at the synapse. Acta Neuropathol Commun. 2019;7(1):109.

15. Casaletto KB, Nichols E, Aslanyan V, Simone SM, Rabin JS, LaJoie R, et al. Sex-specific effects of microglial activation on Alzheimer’s disease proteinopathy in older adults. Brain. 2022.

16. Xiang X, Wind K, Wiedemann T, Blume T, Shi Y, Briel N, et al. Microglial activation states drive glucose uptake and FDG-PET alterations in neurodegenerative diseases. Sci Transl Med. 2021;13(615):eabe5640.

17. Oh H, Madison C, Baker S, Rabinovici G, Jagust W. Dynamic relationships between age, amyloid-beta deposition, and glucose metabolism link to the regional vulnerability to Alzheimer’s disease. Brain. 2016;139(Pt 8):2275–89.

18. Gordon BA, Blazey TM, Su Y, Hari-Raj A, Dincer A, Flores S, et al. Spatial patterns of neuroimaging biomarker change in individuals from families with autosomal dominant Alzheimer’s disease: a longitudinal study. Lancet Neurol. 2018;17(3):241–50.

19. Ashraf A, Fan Z, Brooks DJ, Edison P. Cortical hypermetabolism in MCI subjects: a compensatory mechanism? Eur J Nucl Med Mol Imaging. 2015;42(3):447–58.

20. Arenaza-Urquijo EM, Bejanin A, Gonneaud J, Wirth M, La Joie R, Mutlu J, et al. Association between educational attainment and amyloid deposition across the spectrum from normal cognition to dementia: neuroimaging evidence for protection and compensation. Neurobiology of aging. 2017;59:72–9.

21. Ewers M, Biechele G, Suarez-Calvet M, Sacher C, Blume T, Morenas-Rodriguez E, et al. Higher CSF sTREM2 and microglia activation are associated with slower rates of beta-amyloid accumulation. EMBO Mol Med. 2020;12(9):e12308.

22. Ewers M, Franzmeier N, Suarez-Calvet M, Morenas-Rodriguez E, Caballero MAA, Kleinberger G, et al. Increased soluble TREM2 in cerebrospinal fluid is associated with reduced cognitive and clinical decline in Alzheimer’s disease. Sci Transl Med. 2019;11(507).

23. Morenas-Rodriguez E, Li Y, Nuscher B, Franzmeier N, Xiong C, Suarez-Calvet M, et al. Soluble TREM2 in CSF and its association with other biomarkers and cognition in autosomal-dominant Alzheimer’s disease: a longitudinal observational study. Lancet Neurol. 2022;21(4):329–41.

24. Parhizkar S, Arzberger T, Brendel M, Kleinberger G, Deussing M, Focke C, et al. Loss of TREM2 function increases amyloid seeding but reduces plaque-associated ApoE. Nat Neurosci. 2019;22(2):191–204.

25. Keren-Shaul H, Spinrad A, Weiner A, Matcovitch-Natan O, Dvir-Szternfeld R, Ulland TK, et al. A Unique Microglia Type Associated with Restricting Development of Alzheimer’s Disease. Cell. 2017;169(7):1276–90 e17.

26. Krasemann S, Madore C, Cialic R, Baufeld C, Calcagno N, El Fatimy R, et al. The TREM2-APOE Pathway Drives the Transcriptional Phenotype of Dysfunctional Microglia in Neurodegenerative Diseases. Immunity. 2017;47(3):566–81 e9.

27. Ewers M, Franzmeier N, Surez-Calvet M, Morenas-Rodriguez E, Caballero MAA, Kleinberger G, et al. Increased soluble TREM2 in cerebrospinal fluid is associated with reduced cognitive and clinical decline in Alzheimer’s disease. Science Translational Medicine. 2019;11(507).

28. Franzmeier N, Suarez-Calvet M, Frontzkowski L, Moore A, Hohman TJ, Morenas-Rodriguez E, et al. Higher CSF sTREM2 attenuates ApoE4-related risk for cognitive decline and neurodegeneration. Mol Neurodegener. 2020;15(1):57.

29. Suarez-Calvet M, Kleinberger G, Araque Caballero MA, Brendel M, Rominger A, Alcolea D, et al. sTREM2 cerebrospinal fluid levels are a potential biomarker for microglia activity in early-stage Alzheimer’s disease and associate with neuronal injury markers. EMBO Mol Med. 2016;8(5):466–76.

30. Suarez-Calvet M, Morenas-Rodriguez E, Kleinberger G, Schlepckow K, Araque Caballero MA, Franzmeier N, et al. Early increase of CSF sTREM2 in Alzheimer’s disease is associated with tau related-neurodegeneration but not with amyloid-beta pathology. Mol Neurodegener. 2019;14(1):1.

31. Palmqvist S, Scholl M, Strandberg O, Mattsson N, Stomrud E, Zetterberg H, et al. Earliest accumulation of beta-amyloid occurs within the default-mode network and concurrently affects brain connectivity. Nat Commun. 2017;8(1):1214.

32. Landau SM, Mintun MA, Joshi AD, Koeppe RA, Petersen RC, Aisen PS, et al. Amyloid deposition, hypometabolism, and longitudinal cognitive decline. Ann Neurol. 2012;72(4):578–86.

33. Royse SK, Minhas DS, Lopresti BJ, Murphy A, Ward T, Koeppe RA, et al. Validation of amyloid PET positivity thresholds in centiloids: a multisite PET study approach. Alzheimers Res Ther. 2021;13(1):99.

34. Kleinberger G, Yamanishi Y, Suarez-Calvet M, Czirr E, Lohmann E, Cuyvers E, et al. TREM2 mutations implicated in neurodegeneration impair cell surface transport and phagocytosis. Sci Transl Med. 2014;6(243):243ra86.

35. Avants BB, Tustison NJ, Song G, Cook PA, Klein A, Gee JC. A reproducible evaluation of ANTs similarity metric performance in brain image registration. Neuroimage. 2011;54(3):2033–44.

36. Klunk WE, Koeppe RA, Price JC, Benzinger TL, Devous MD, Sr., Jagust WJ, et al. The Centiloid Project: standardizing quantitative amyloid plaque estimation by PET. Alzheimers Dement. 2015;11(1):1–15 e1-4.

37. R Core Team. R: A language and environment for statistical computing. Vienna, Austria: R Foundation for Statistical Computing; 2021.

38. Landau SM, Harvey D, Madison CM, Koeppe RA, Reiman EM, Foster NL, et al. Associations between cognitive, functional, and FDG-PET measures of decline in AD and MCI. Neurobiol Aging. 2011;32(7):1207–18.

39. Schaefer A, Kong R, Gordon EM, Laumann TO, Zuo XN, Holmes AJ, et al. Local-Global Parcellation of the Human Cerebral Cortex from Intrinsic Functional Connectivity MRI. Cereb Cortex. 2018;28(9):3095–114.

40. Palmqvist S, Mattsson N, Hansson O, Alzheimer’s Disease Neuroimaging I. Cerebrospinal fluid analysis detects cerebral amyloid-beta accumulation earlier than positron emission tomography. Brain. 2016;139(Pt 4):1226–36.

41. Ossenkoppele R, Reimand J, Smith R, Leuzy A, Strandberg O, Palmqvist S, et al. Tau PET correlates with different Alzheimer’s disease-related features compared to CSF and plasma p-tau biomarkers. EMBO Mol Med. 2021;13(8):e14398.

42. Bhaskar K, Konerth M, Kokiko-Cochran ON, Cardona A, Ransohoff RM, Lamb BT. Regulation of tau pathology by the microglial fractalkine receptor. Neuron. 2010;68(1):19–31.

43. Lee S, Xu G, Jay TR, Bhatta S, Kim KW, Jung S, et al. Opposing effects of membrane-anchored CX3CL1 on amyloid and tau pathologies via the p38 MAPK pathway. The Journal of neuroscience : the official journal of the Society for Neuroscience. 2014;34(37):12538–46.

44. Ising C, Venegas C, Zhang S, Scheiblich H, Schmidt SV, Vieira-Saecker A, et al. NLRP3 inflammasome activation drives tau pathology. Nature. 2019;575(7784):669–73.

45. Stozicka Z, Zilka N, Novak P, Kovacech B, Bugos O, Novak M. Genetic background modifies neurodegeneration and neuroinflammation driven by misfolded human tau protein in rat model of tauopathy: implication for immunomodulatory approach to Alzheimer’s disease. J Neuroinflammation. 2010;7:64.

46. Hopp SC, Lin Y, Oakley D, Roe AD, DeVos SL, Hanlon D, et al. The role of microglia in processing and spreading of bioactive tau seeds in Alzheimer’s disease. J Neuroinflammation. 2018;15(1):269.

47. Cheng-Hathaway PJ, Reed-Geaghan EG, Jay TR, Casali BT, Bemiller SM, Puntambekar SS, et al. The Trem2 R47H variant confers loss-of-function-like phenotypes in Alzheimer’s disease. Mol Neurodegener. 2018;13(1):29.

48. Jin SC, Benitez BA, Karch CM, Cooper B, Skorupa T, Carrell D, et al. Coding variants in TREM2 increase risk for Alzheimer’s disease. Hum Mol Genet. 2014;23(21):5838–46.

49. Leyns CEG, Gratuze M, Narasimhan S, Jain N, Koscal LJ, Jiang H, et al. TREM2 function impedes tau seeding in neuritic plaques. Nat Neurosci. 2019;22(8):1217–22.

50. Gratuze M, Chen Y, Parhizkar S, Jain N, Strickland MR, Serrano JR, et al. Activated microglia mitigate Abeta-associated tau seeding and spreading. J Exp Med. 2021;218(8).

51. Hansson O. Biomarkers for neurodegenerative diseases. Nat Med. 2021;27(6):954–63.

52. Jay TR, Hirsch AM, Broihier ML, Miller CM, Neilson LE, Ransohoff RM, et al. Disease Progression-Dependent Effects of TREM2 Deficiency in a Mouse Model of Alzheimer’s Disease. J Neurosci. 2017;37(3):637–47.

53. Benzinger TL, Blazey T, Jack CR, Jr., Koeppe RA, Su Y, Xiong C, et al. Regional variability of imaging biomarkers in autosomal dominant Alzheimer’s disease. Proc Natl Acad Sci U S A. 2013;110(47):E4502–9.

54. Salvado G, Mila-Aloma M, Shekari M, Ashton NJ, Operto G, Falcon C, et al. Reactive astrogliosis is associated with higher cerebral glucose consumption in the early Alzheimer’s continuum. Eur J Nucl Med Mol Imaging. 2022.

55. Knopman DS, Jones DT, Greicius MD. Failure to demonstrate efficacy of aducanumab: An analysis of the EMERGE and ENGAGE trials as reported by Biogen, December 2019. Alzheimers Dement. 2021;17(4):696–701.

56. Fleisher AS, Chen K, Quiroz YT, Jakimovich LJ, Gutierrez Gomez M, Langois CM, et al. Associations between biomarkers and age in the presenilin 1 E280A autosomal dominant Alzheimer disease kindred: a cross-sectional study. JAMA Neurol. 2015;72(3):316–24.

57. Jack CR, Jr., Bennett DA, Blennow K, Carrillo MC, Dunn B, Haeberlein SB, et al. NIA-AA Research Framework: Toward a biological definition of Alzheimer’s disease. Alzheimers Dement. 2018;14(4):535–62.

58. Bennett DA, Schneider JA, Wilson RS, Bienias JL, Arnold SE. Neurofibrillary tangles mediate the association of amyloid load with clinical Alzheimer disease and level of cognitive function. Arch Neurol. 2004;61(3):378–84.

59. Wang L, Benzinger TL, Su Y, Christensen J, Friedrichsen K, Aldea P, et al. Evaluation of Tau Imaging in Staging Alzheimer Disease and Revealing Interactions Between beta-Amyloid and Tauopathy. JAMA Neurol. 2016;73(9):1070–7.

60. Ossenkoppele R, Schonhaut DR, Scholl M, Lockhart SN, Ayakta N, Baker SL, et al. Tau PET patterns mirror clinical and neuroanatomical variability in Alzheimer’s disease. Brain. 2016;139(Pt 5):1551–67.

61. Jack CR, Jr., Knopman DS, Jagust WJ, Petersen RC, Weiner MW, Aisen PS, et al. Tracking pathophysiological processes in Alzheimer’s disease: an updated hypothetical model of dynamic biomarkers. Lancet Neurol. 2013;12(2):207–16.

62. Schindler SE, Cruchaga C, Joseph A, McCue L, Farias FHG, Wilkins CH, et al. African Americans Have Differences in CSF Soluble TREM2 and Associated Genetic Variants. Neurol Genet. 2021;7(2):e571.

